# Cohort study of fever neonates with confirmed community infection of SARS-CoV-2 Omicron: A study Protocol

**DOI:** 10.1101/2022.12.28.22283987

**Authors:** Jicheng Li, Wenwen He, Yujie Qi, Yao Meng, Xiaoxia Peng, Mingyan Hei

## Abstract

**Introduction:** Reports on neonatal coronavirus disease (COVID) have been focused on vertical transmission. There was little information on prevalence of neonates with COVID till up to 18 months of age. It is well known that breastfeeding is beneficial for the growth and development of infants. We hypothesized that breastfeeding will be beneficial for a better prevalence of neonatal COVID. The objective of this study is to explore the prognosis of fever neonates with confirmed community infection of SARS-CoV-2 Omicron, and to clarify whether human milk breastfeeding is beneficial for reducing the rate of severe SARS-CoV-2 Omicron infection in neonates.

**Methods and analysis:** This is a prospective single centre cohort study. Study period is from December, 2022 to December 2024. Inclusion criteria are: (1) Age ≤ 28 days or corrected age ≤ 44 weeks. (2) Fever. (3) Both tests (throat swab) of nucleic acid and antigen of SARS-CoV-2 Omicron were positive. (4) Parents signed the consents. Exclusion criteria is confirmed brain malformations. Patients will be classified into breastfeeding, mixed feeding, and formula feeding groups. The estimate sample size will be 200. The throat swab of infants will be collected for SARS-CoV-2 omicron nucleic acid ad antigen examination. Neonatal COIVD patients will be treated in the Out-Patient Department or admitted to Neonatal Intensive Care Unit according to the severity of infection. All patients will be followed at 3/6/12/18M of age. The endpoint to study was at 18 months of age. Data will be collected by Case Record Form and Electronic Data Capture from the History of In-hospital System. The primary outcome was the rate of severe SARS-CoV-2 infection. *SPSS 20.0* software will used for statistical analysis.

**Ethics and dissemination:** It is approved by local Institute of Ethics Review Board (#[2022]-E-240-Y).

**Registration:** It is registered in the Chinese Clinical Trail Registry (http://www.chictr.org.cn) (ChiCTR2200067148)

**Strengths and limitations of this study:**

1. This is a prospective single centre cohort study for fever neonates with confirmed community infection of SARS-CoV-2 Omicron.
2. Patients are categorized into breastfeeding group, mixed feeding group, and formula group.
3. The primary outcome is rate of severe SARS-CoV-2 infection.
4. The secondary outcomes include in-hospital outcomes and follow-up outcomes at 3M/6M/12//18M.
5. Limitation are the observational nature and the single centre data.

## Introduction

The global pandemic outbreak of coronavirus disease (COVID) was started in 2019. Till now, the number of neonatal COVID has been relatively less than adults. Reports on neonates have been focused on perinatal vertical transmission^[1][2]^, as well as the safety issues of breastfeeding for neonates^[3]^. There were a few publications reported the late onset of infection of SARS-CoV-2 in neonates^[4][5]^, among which, neonates presented mild fever, mild sniff nose or running nose, or even asymptomatic. The latest report from Shanghai China was published in *Chinese Pediatric Journal* in December 2022, in which 16 COVID neonates were included, and the clinical characteristics and discharge outcomes were described^[6]^. However, in the report there were only 11 neonates presented fever and the highest temperature was 38.4°C. So far, there was little information reported on the prevalence of neonates with confirmed community infection of SARS-CoV-2 between birth to 18 months of age.

SARS-CoV-2 is easy to have variant strains, causing its contagiosity and pathogenicity variable^[7]^. In December 2022, there were a large amount of population infected with SARS-CoV-2 in Beijing, and the strain was Omicron BF7 according to the surveillance by Chinese Centers for Disease Control (CDC)^[8]^. Most people were community infected and presented with high fever, severe sore throat, fatigue, body ache, and coughing. Meanwhile, there was a sharply increased number of community infected neonatal patients. High fever, crying, and poor feeding were the commonest symptoms of newborn COVID patients in Beijing during this period, some of the newborns need to be admitted to hospitals. Due to limited information about the community infection of SARS-CoV-2 Omicron in neonates who have apparent infection symptoms, both medical professionals and parents wanted to know: Whether the SARS-CoV-2 Omicron infection at neonatal period will cause negative influence to the infants’ growth and development? Is there any protective factor which may reduce the rate of severe SARS-CoV-2 Omicron cases?

It is well known that breastfeeding is beneficial for the growth and development of infants. We hypothesized that breastfeeding will be beneficial for a better prevalence from birth to 18 months old when newborns get infected with SARS-CoV-2. The objective of this study is **t**o explore the prognosis of fever neonates with confirmed community infection of SARS-CoV-2 Omicron, and to clarify whether human milk breastfeeding is beneficial to reduce the rate of severe SARS-CoV-2 Omicron infection in neonates.

## Methods and analysis

### Study design

This is a prospective single centre cohort study.

### Settings and Participants

The study will be completed in Neonatal Center, Beijing Children’s Hospital, Capital Medica University, Beijing, China. The study period is from December 25, 2022 to December 31, 2024. Patients diagnosed from December 2022 to May 2023 will be approached. All enrolled patients will be followed up from December 2022 to December 2023. The endpoint of study is 18 months of age.

Infants who meet all of the following 5 points will be included: (1) Infants who went to Beijing Children’s Hospital between December, 2022 to May, 2023. (2) Age ≤ 28 days or corrected age ≤ 44 weeks. (3) Fever. (4) Both tests (throat swab) of nucleic acid and antigen of SARS-CoV-2 Omicron were positive. (5) Parents signed the consents.

Infants who had confirmed brain malformations will be excluded.

All enrolled patients will be categorized into 3 groups according to their previous feeding: Breastfeeding group, mixed feeding group, formula group.

### Sample size

Approximately 200 neonatal patients of SARS-CoV-2 Omicron infection will be enrolled, with a minimal patient number of 30 in each of the three groups. Technically, all patients who meet the inclusion and exclusion criteria in our hospital between December 25, 2022 to May 30, 2024 will be approached, so that the last patient enrolled can finish his/her 18M follow-up in the Out-patient Clinic.

### Variables

The primary outcome is rate of severe SARS-CoV-2 infection.

The secondary outcomes are in-hospital outcomes (rate of negative nucleic acid test of SARS-CoV-2 on day 7, time of negative nucleic acid test of SARS-CoV-2, time of negative antigen test of SARS-CoV-2, persistent time of fever, length of stay in NICU, medical expenses, mortality rate) and follow-up outcomes after discharge home (weight/head circumference/height at 3M/6M/12//18M, development quotient (DQ) at 3M/6M/12//18M,head magnetic resonance imaging at 3M/12M).

The potential confounders are male gender, age of onset, and number of family members with confirmed COIVD meanwhile.

### Definitions

Fever refers to the rectal temperature is ≥38°C^[9][10]^.

Community infection of SARS-CoV-2 Omicron of neonates refers to the neonate who meet the following criteria: (1) His/her age is older than 7 days, and (2) home care, and (3) his/her throat swab examination of both nucleic acid and antigen of SARS-CoV-2 Omicron are positive, and (4) more than 1 family members have been confirmed to be COVID patient meanwhile.

Severe case of neonatal SARS-CoV-2 Omicron infection refers to neonates who meet one of the following criteria^[4][5][11]^: (1) Requiring invasive or non-invasive ventilation, or (2) symptoms and signs of septic shock (such as cyanosis or mosaic on skin, lethargy, cold extremities, no eating), or (3) convulsion, or (4) cyanosis with shallow breathing/breathing difficulty/grunting/apnea, or (5) evidences of bone marrow arrest (severe anemia, and/or white blood cell counting <3000×10^9^/L, and/or platelet counting <100×10^12^/L), or (6) severe stress hyperglycemia.

Breastfeeding refers to the infants’ only food is his/her mother’s breast milk (either fed by bottle or by putting on the breasts) and has never been fed with any formula.

Mixed feeding refers to the infants are fed by his/her mother’s own breast milk and formula.

### Clinical management of SARS-CoV-2 Omicron infection in neonates

Patients will be diagnosed and managed by neonatologists at Pediatric Out-Patient Department (OPD) or by pediatricians at Emergency Department of Beijing Children’s Hospital. Patients will be treated and managed either in OPD or being admitted to NICU, mainly according to patients’ severity. All patients will complete the throat swab examination of both nucleic acid and antigen of SARS-CoV-2 Omicron at OPD.

Indications for OPD management are: (1) The patient’ symptom is only mild fever and persists for less than 24h hours, and (2) his/her general condition is good and feeding is normal, and (3) Whole blood counting and C-reaction protein are normal, and (4) parents are confident to take care of the infant. OPD treatments are mainly physical methods of decreasing body temperature (such as bathing, massage, mild cooling). Parents will be educated to observe the infants’ general condition and skin color, as well as extra-supplementation of water feeding during fever.

Indications of being admitted to NICU are: (1) Confirmed or suspected severe case of neonatal SARS-CoV-2 Omicron infection, or (2) Higher fever (body temperature ≥39°C), especially persisted for longer than 24 hours, or (3) Infants presents recurrent vomiting or diarrhea. Patients will be kept in isolation area. Treatments in NICU are mainly physical methods of decreasing body temperature, nursing and vital sings monitoring, bottle feeding (according to parents’ own decision whether to continue feed with his/her mother’s own milk or formula. Parents need to sign a consent for formula feeding). Enteral feeding is encouraged, but intravenous fluid will be given according to patient’s condition in order to maintain a balanced body fluid in/out. On day 5 after admission, throat swab examination of both nucleic acid and antigen of SARS-CoV-2 Omicron will be repeated every 2 days (in a 24-hour-interval). Other clinical managements are adjustable in an individualized principal.

Discharge criteria are: (1) The infants’ general condition is good, and meet the general discharge criteria for newborns (including vital signs have been stable and body temperature has been normal for more than 24 hours, and well tolerate enteral feeding)^[11]^. (2) Parents are confident to take care of the infants^[12]^. (3) There is at least 1 negative result of nucleic acid examination of SARS-CoV-2 Omicron.

After discharge, all patients will be followed in the follow-up clinic at 3M, 6M, 12M and 18 M of age by pediatricians. Growth assessment of weight/head circumference/height will be recorded, as well as the developmental assessment by either Bayley assessment or other development assessment tools.

### Examination of SARS-CoV-2 Omicron

Registered nurses will be in charge of the throat swab and blood collection, and neonatologists will be in charge of cerebral spinal fluid (if needed). All samples will be sent to the virus lab for nucleic acid examination of SARS-CoV-2 Omicron by PCR method within 2 hours. The antigen examination of SARS-CoV-2 Omicron will be completed according to the kit instruction by aerosol method right after the throat swab was collected.

The positive result of nucleic acid examination will be judged only after 2 times of nucleic acid examination were both positive (one kit is from *Da-an Company*, and another kit is from *Zhuo-cheng-hui-sheng Company*, Beijing, China). The positive result of antigen examination of SARS-CoV-2 Omicron is 2 bands on the cartridge within 15 minutes (*Lot.#20223400365*) (the kit is from *Zhong-yuan-hui-ji Ltd. Company*, Chongqing, China).

### Data source and measurements

A designated researcher will be assigned to collect and manage the data by using 2 methods: (1) Case Record Form (CRF), and (2) Electronic Data Capture (EDC) from the History of In-hospital System (HIS).

Demographic data (gestational age, weight, gender, maternal disease, feeding, admission weight, discharge weight) and baseline data of disease (age at onset, body temperature on admission, respiratory symptoms, whole blood count, blood culture, CSF examination, nucleic acid test of SARS-CoV-2 on admission, antigen test of SARS-CoV-2 on admission) will be collected.

### Statistical analysis

The Shapiro-Wilk test was firstly used to explore whether the data were normally distributed. Data with normal distribution were expressed as (mean ± standard deviation), while data with non-normal distribution were expressed as medium (IQR: Q25-Q75). Two-sample *t* tests or rank sum tests were used to compare the continuous variables (e.g., changes in birth weight) for two independent variables. The *chi-square* test was used to compare dichotomous variables. To further compare the rate of severe SARS-CoV-2 infection between groups, we used a generalized linear model to adjust for potential confounders identified from descriptive comparisons of baseline characteristics. Subgroup analysis was also performed. The potential confounders included male gender, number of family members with confirmed COIVD meanwhile. Tests with values of *P* < 0.05 were considered significant. There will be no sensitivity analysis. Statistical analyses will be performed using *IBM SPSS 20.0* software (version 26); the figure will be drawn using GraphPad Prism (version 9).

## Monitoring and Evaluation of the Study

The aim of monitoring and evaluation is to reduce the bias of the study. The major bias is the selective bias due to the nature of single centre cohort study. The monitoring and evaluation of the study are composed of the following 3 sections:

### Monitoring of research process

A designated research coordinator will be in charge of coordinating the research process including interacting with different departments to ensure program implementation and data collection. The principal investigator will summarize the collected data, analyze the data, find problems and make decisions regarding how to solve the problems.

### Quality control procedures

The quality control procedures include: (1) Carefully train participating researchers in the study before enrolling patients. (2) To designate a full-time research coordinator to oversee project, and to designated full time graduate students to collect the data. (3) PI will routinely work on the analysis of enrollment and completion numbers and feedback from participating researchers, work with statistical specialists and discuss about the progress of the study.

### Procedures to ensure compliance with ethical standards

The procedures include: (1) To obtain ethic approval from the Ethics Committee for Clinical Research in Beijing Children’s Hospital. (2) To obtain informed consent from each participating parent before joining the study. (3) To setup a system for incidence reporting, including the use of time limitations (less than 24 hours) and the need for a formal written report. (4) To carefully conduct the study based on procedure in the protocol. (5) To use patient numbers instead of patient names when summarizing the data. (6) To get parents’ written approval if their names and pictures are going to appear on the research final report or publications. (7) Respect parents’ decision to join or leave the study at any time they wish.

## Data Availability

All data produced in the present study are available upon reasonable request to the authors

## Abbreviations

CDC: Centers for Disease Control
COVID: coronavirus disease
DQ: development quotient
IRB: Institutional Ethics Review Board
NICU: neonatal intensive care unit
OPD: out-patient department

## Ethics and dissemination

The study was approved by the Institute of Ethics Review Board (IRB) of Beijing Children’s Hospital (#[2022]-E-240-Y). Data will be deposit in both Department of Neonatology and the Institutional Review Board, which will not be shared with public. Results will be disseminated by publications as soon as possible.

## Author Contributions

(I) Conception and design: JL, PX, MH; (II) Administrative support: MH; (III) Provision of study materials or patients: JL, WH, YM, YQ; (IV) Collection and assembly of data: JL, WH, YM, YQ, MY; (V) Data analysis and interpretation: JL, WH, YQ, PX, MY; (VI) Manuscript writing: All authors; (VII) Final approval of manuscript: All authors.

